# COVID-19 management in a UK NHS Foundation Trust with a High Consequence Infectious Diseases centre: a detailed descriptive analysis

**DOI:** 10.1101/2020.05.14.20100834

**Authors:** Kenneth F. Baker, Aidan T. Hanrath, Ina Schim van der Loeff, Su Ann Tee, Richard Capstick, Gabriella Marchitelli, Ang Li, Andrew Barr, Alsafi Eid, Sajeel Ahmed, Dalvir Bajwa, Omer Mohammed, Neil Alderson, Clare Lendrem, Dennis Lendrem, COVID-19 Control Group, COVID-19 Clinical Group, Lucia Pareja-Cebrian, Andrew Welch, Joanne Field, Brendan A.I. Payne, Yusri Taha, David A. Price, Christopher Gibbins, Matthias L. Schmid, Ewan Hunter, Christopher J.A. Duncan

**Affiliations:** Translational and Clinical Research Institute, Newcastle University, UK; The Newcastle upon Tyne Hospitals NHS Foundation Trust, Newcastle upon Tyne, UK; NIHR In Vitro Diagnostics Cooperative, Newcastle University, UK; National Institute of Health Research (NIHR) Biomedical Research Centre, Newcastle University, UK; NUTH COVID control group: Margaret Gray (Deputy COO, Chair), Derna Campbell, Michael Clark, Angela Cobb, Sue Cook, Melanie Cunningham, Lewis Gibson, Lisa Guthrie, Elizabeth Harris, David Kinnersley, Allison Sykes, Michael Wright; NUTH COVID clinical group: Kevin Brennan, Graham Burns, Matthew Cadamy, Ian Clement, Jennifer Collins, Jill Dixon, Martin Duddy, Adam Evans, Simon Hill, Kelly Hunt, Simon Kerr, Stuart Little, Christopher Mountford, Anne Pelham, Sarah Platt, Ulrich Schwab, Julie Samuel, Ally Speight, Nadia Stock, Jason Urron, Jon Walton, Sophie West.

## Abstract

**Background:** Recent large national and international cohorts describe the baseline characteristics and outcome of hospitalised patients with COVID-19, however there is little granularity to these reports. We aimed to provide a detailed description of a UK COVID-19 cohort, focusing on clinical decisions and patient journeys.

**Methods:** We retrospectively analysed the management and 28-day outcomes of 316 consecutive adult patients with SARS-CoV-2 PCR-confirmed COVID-19 admitted to a large NHS Foundation Trust with a tertiary High Consequence Infectious Diseases centre in the North of England.

**Findings:** Most patients were elderly (median age 75) with multiple comorbidities. One quarter were admitted from residential or nursing care. Symptoms were consistent with COVID-19, with cough, fever and/or breathlessness in 90.5% of patients. Two thirds of patients had severe disease on admission. Mortality was 81/291 (27.8%). Most deaths were anticipated; decisions to initiate respiratory support were individualised after consideration of patient wishes, premorbid frailty and comorbidities, with specialist palliative care input where appropriate. 22/291 (7.6%) patients were intubated and 11/22 (50%) survived beyond discharge. Multiple logistic regression identified age as the most significant risk factor for death (OR 1.09 [95% CI 1.06 - 1.12] per year increase, p < 0.001).

**Interpretation:** These findings provide important clinical context to outcome data. Deaths were anticipated, occurring in patients with advance decisions on ceilings of treatment. Age was the most significant risk factor for death, confirming that demographic factors in the population are a major influence on hospital mortality rates.

**Funding:** Funding was not required.

## Introduction

The first two patients with COVID-19 in the United Kingdom (UK) received inpatient care at The Newcastle upon Tyne Hospitals NHS Foundation Trust, one of five airborne High Consequence Infectious Diseases centres, on 31 January 2020.1 Since then, 211,364 patients have tested positive for SARS-CoV-2 in the UK and 32,692 people have died of COVID-19 (as of 11 May 2020),2 putting the UK second only to the USA in total deaths. Early studies from China,3 Spain,4 and the USA5 reported mortality rates of 20.7 - 28.3% of hospitalised patients, but differences in population demographics, health behaviours, and systems of healthcare between these countries may influence both outcome and how outcomes are recorded.

The characteristics of 16,749 patients with COVID-19 managed in UK National Health Service (NHS) hospitals were recently reported by the International Severe Acute Respiratory and emerging Infections Consortium (ISARIC).6 Overall mortality was 33%, and within that cohort patients who died were older and carried a high burden of comorbidity. Across the UK, clinical decisions about appropriate ceilings of treatment at the individual patient level are made daily, however information regarding this key aspect of management is lacking in the ISARIC dataset. Such decisions may impact death rates and influence our understanding of risk factors associated with outcome in COVID-19. We sought to provide a comprehensive description of a UK COVID-19 inpatient cohort, focusing on clinical management pathways and outcome.

## Methods

The Newcastle upon Tyne Hospitals NHS Foundation Trust (NUTH) is a large tertiary academic medical centre serving the population of Newcastle upon Tyne (estimated 302,820)7 and the wider North East of England. NUTH is one of five designated centres for airborne High Consequence Infectious Diseases in the UK. Following isolation of confirmed cases during the initial ‘containment’ phase of the UK response (until 12th March), emergency and inpatient care was provided to patients presenting with symptoms of COVID-19 during the ongoing community transmission phase of the epidemic. In accordance with NHS England guidelines, combined nose and throat swabs and/or sputum samples were obtained for SARS-CoV-2 reverse-transcriptase polymerase chain reaction (PCR) for all patients admitted to hospital who met clinical criteria. Initially PCR testing was performed using the Public Health England RdRp assay until 7th April, followed by the Altona Diagnostics (from 1st April) and Roche cobas 6800 (from 7th April) assay platforms.

We searched our electronic health records to identify all consecutive patients admitted to NUTH between 8^th^ January to ^16^ April 2020 inclusive with a positive SARS-CoV-2 PCR result. Of 362 patients identified, 46 were excluded from analysis: 14 had no relevant admission at the time of testing, 21 were already inpatients at the time of infection, six were below 18 years of age, and five were asymptomatic patients screened on admission to hospital for an unrelated reason. Six patients were inpatient transfers with COVID-19 for whom first recorded observations were not available.

The study was registered as a clinical service evaluation with The Newcastle upon Tyne Hospitals NHS Foundation Trust and was exempt from ethical approval,with analysis of anonymised healthcare data approved by the Caldicott Guardian. Electronic health records were retrospectively reviewed by a team of medical doctors with the aid of a standardised version-controlled data collection template (Excel, Microsoft Corporation) with internal data validation restrictions. Data included demographic details, presenting symptoms, and baseline clinical and laboratory parameters. Comorbidities and Clinical Frailty Scale8 were defined as clinician reported. Radiological findings were classified according to the British Society of Thoracic Imaging criteria,9 as documented by the reporting radiologist. Clinical status (death, invasive ventilation, non-invasive pressure support including continuous positive airway pressure and bilevel positive airway pressure, oxygen therapy, or discharged alive) as per the World Health Organisation (WHO) ordinal scale for COVID-1910 was recorded for each calendar day for a total of 28 days after hospital admission, or until a censor date of 5^th^ May 2020. All inpatient and outpatient deaths occurring up to this censor date were also recorded. Daily system updates allow us to record deaths in the community via reporting through primary care, and thus we assumed patients were alive at home unless they were flagged as an inpatient or deceased on our records. Collected data were merged and reviewed for errors (0.8%) and missing data (ATH, ISvdL, and KFB) prior to analysis.

Analyses were performed in R (version 3.6.0, R Core Team) and SAS JMP Pro(version 13.2.1, SAS Institute, Cary NC). The significance of departure of observed male sex proportion from an expected value of 0.5 was assessed using the one sample z-test. Tests of differences in proportions γ^2^(x test) and continuous data (Wilcoxon rank sum test) were performed between contrast groups where stated. Odds ratios for death were calculated between death and survival groups by univariate and multiple logistic regression, with a two-tailed α <0.05 considered statistically significant.

## Results

### Clinical features at presentation

A total of 316 patients were identified with a median (IQR) [range] age of 75 (60 – 83) [23 – 101] years. 281/303 (92.7%) patients were white British, white Irish or other white ethnicity. Over half the cohort (54.7%) was male. Males were disproportionately represented at all ages under 70 years (75/124 [60.5%], p = 0.02). Interestingly, broadly similar proportions of men and women were admitted in those aged 70 years and over (98/192 [51.0%] male, p = 0.51). Twenty seven of 316 (8.5%) patients were healthcare workers.

Median (IQR) [range] symptom duration prior to admission was 5 days (2-9) [0-42]. The most common presenting symptoms were cough (224 [70.9%]), fever (211 [66.8%]), and breathlessness (197 [62.3%]), with 286 (90.5%) patients presenting with at least one of these symptoms. Many patients also reported fatigue (128 [40.5%]), myalgia and/or arthralgia (72 [22.8%]), and diarrhoea (64 [20.3%]). 60 (19.0%) patients were admitted from a residential or nursing home. 69 (21.8%) patients had been treated for their symptoms in primary care prior to admission, of which 48/69 (69.6%) had received a course of oral antibiotics. 253/316 (80.1%) patients had at least one major comorbidity (Table 1), the most common of which were hypertension (133 [42.1%]), chronic kidney disease (77 [24.4%]), ischaemic heart disease (65 [20.6%]), and dementia (55 [17.4%]).

**Table 1.** Cohort characteristics including univariate logistic regression analyses and associated p values. Odds ratios are stated per 1 unit increase in the independentvariable unless otherwise stated.

|  | N | Cohort (316) | Survived (210) | Died (81) | Univariate OR<sub>death</sub> (95% CI) | p |
| --- | --- | --- | --- | --- | --- | --- |
| Age (years) | 316 | 75 (60 – 83) | 68 (55 – 80) | 82 (76 – 89) | 1.09 (1.06 - 1.12) | <0.001 |
| Male sex | 316 | 173 (54.7) | 113 (53.8) | 46 (56.8) | 1.13 (0.67 - 1.9) | 0.647 |
| White ethnicity | 303 | 281/303 (92.7) | 185/203 (91.1) | 74/76 (97.4) | 3.6 (1.01 – 23.0) | 0.091 |
| Symptom duration (days) | 316 | 5 (2 – 9) | 6 (3 – 9) | 3 (1 – 7) | 0.92 (0.86 - 0.97) | 0.004 |
| Fever | 316 | 211 (66.8) | 150 (71.4) | 45 (55.6) | 0.5 (0.29 - 0.85) | 0.010 |
| Cough | 316 | 224 (70.9) | 157 (74.8) | 52 (64.2) | 0.61 (0.35 - 1.06) | 0.074 |
| Sputum | 316 | 76 (24.1) | 56 (26.7) | 13 (16.0) | 0.53 (0.26 – 1.00) | 0.059 |
| Breathlessness | 316 | 197 (62.3) | 126 (60.0) | 58 (71.6) | 1.68 (0.97 - 2.97) | 0.067 |
| Fatigue | 316 | 128 (40.5) | 91 (43.3) | 31 (38.3) | 0.81 (0.48 - 1.36) | 0.433 |
| Myalgia/arthralgia | 316 | 70 (22.2) | 57 (27.1) | 11 (13.6) | 0.42 (0.2 - 0.83) | 0.016 |
| Diarrhoea | 316 | 64 (20.3) | 48 (22.9) | 12 (14.8) | 0.59 (0.28 - 1.14) | 0.132 |
| Any comorbidity<sup>1</sup> | 316 | 250 (79.1) | 157 (74.8) | 75 (92.6) | 4.22 (1.87 - 11.4) | 0.001 |
| Respiratory comorbidity<sup>2</sup> | 316 | 101 (32.0) | 71 (33.8) | 23 (28.4) | 0.78 (0.44 - 1.35) | 0.377 |
| Heart failure | 316 | 45 (14.2) | 23 (11.0) | 20 (24.7) | 2.67 (1.36 - 5.19) | 0.004 |
| Hypertension | 316 | 133 (42.1) | 77 (36.7) | 43 (53.1) | 1.95 (1.16 - 3.30) | 0.011 |
| Ischaemic heart disease | 316 | 65 (20.6) | 39 (18.6) | 19 (23.5) | 1.34 (0.71 - 2.48) | 0.351 |
| Chronic kidney disease | 316 | 77 (24.4) | 43 (20.5) | 26 (32.1) | 1.84 (1.03 - 3.25) | 0.038 |
| Diabetes mellitus | 316 | 84 (26.6) | 52 (24.8) | 24 (29.6) | 1.28 (0.72 - 2.25) | 0.397 |
| Active cancer | 316 | 33 (10.4) | 22 (10.5) | 10 (12.3) | 1.20 (0.52 - 2.61) | 0.648 |
| Immunosuppression<sup>3</sup> | 316 | 27 (8.5) | 16 (7.6) | 9 (11.1) | 1.52 (0.62 - 3.52) | 0.343 |
| Dementia | 316 | 55 (17.4) | 25 (11.9) | 26 (32.1) | 3.50 (1.87 - 6.58) | <0.001 |
| ACE inhibitor / ARB use | 311 | 78/311 (25.1) | 53/206 (25.7) | 17/80 (21.3) | 0.78 (0.41 - 1.43) | 0.430 |
| Healthcare worker | 316 | 27 (8.5) | 25 (11.9) | 0 (0) | na | 0.983 |
| Admitted from nursing or residential home | 316 | 60 (19.0) | 21 (10.0) | 33 (40.7) | 6.19 (3.32 - 11.8) | <0.001 |
| Severe COVID-19 pneumonia | 308 | 174/308 (56.5%) | 101/207 (48.8) | 56/80 (70.0) | 2.45 (1.43 - 4.30) | 0.001 |
| Haemoglobin (115-165 g/L) | 310 | 134 (115 – 145) | 136 (120 – 145) | 130 (114 – 145) | 0.91 (0.80 - 1.03)<sup>6</sup> | 0.135 |
| Platelets (150-450 x 10<sup>9</sup>/L) | 298 | 217 (174 – 283) | 216 (176 – 280) | 218 (174 – 284) | 1.01 (0.98 - 1.04)<sup>6</sup> | 0.593 |
| Total WBC (4-11 x 10<sup>9</sup>/L) | 311 | 7.20 (5.36 – 9.35) | 6.79 (5.17 – 8.61) | 7.88 (5.85 – 10.43) | 1.00 (0.97 - 1.03) | 0.832 |
| Lymphocytes (1-4 x 10<sup>9</sup>/L) | 311 | 0.88 (0.64 – 1.32) | 0.92 (0.65 – 1.35) | 0.83 (0.62 – 1.33) | 0.99 (0.87 - 1.07) | 0.850 |
| Neutrophils (2-7 x 10<sup>9</sup>/L) | 311 | 5.30 (3.70 – 7.48) | 4.88 (3.54 – 7.04) | 6.60 (4.36 – 9.07) | 1.15 (1.06 - 1.26) | 0.001 |
| eGFR (> 90 mL/min.1.73m<sup>2</sup>) | 307 | 68 (44 – 89) | 76 (54 - >90) | 50 (32 – 73) | 0.87 (0.82 - 0.92)<sup>7</sup> | <0.001 |
| Urea (2.5-7.8 mmol/L) | 308 | 7.0 (4.9 – 11.4) | 6.2 (4.3 – 9.2) | 9.8 (6.5 – 17.0) | 1.11 (1.06 - 1.16) | <0.001 |
| Sodium (133-146 mmol/L) | 308 | 137 (134 – 140) | 137 (134 – 140) | 138 (135 – 143) | 1.07 (1.03 - 1.13) | 0.001 |
| Potassium (3.5-5.3 mmol/L) | 290 | 4.1 (3.8 – 4.5) | 4.1 (3.8 – 4.4) | 4.2 (3.9 – 4.6) | 1.39 (0.93 - 2.12) | 0.113 |
| **Arterial blood pH (7.35-7.45)** | 199 | 7.43 (7.39 – 7.47) | 7.44 (7.4 – 7.48) | 7.40 (7.36 – 7.45) | 0.91 (0.86 - 0.96)<sup>8</sup> | 0.001 |
| **Bicarbonate (mmol/L)** | 198 | 24.8 (22.8 – 26.6) | 25.2 (23.2 – 27.2) | 23.2 (22.0 – 25.5) | 0.86 (0.78 - 0.94) | 0.001 |
| **CRP (<5 mg/L)** | 306 | 72 (30 – 131) | 58 (22 – 118) | 90 (51 – 175) | 1.04 (1.01 - 1.07)<sup>6</sup> | 0.005 |
| **ALT (0-40 U/L)** | 263 | 24 (15 – 39) | 26 (16 – 40) | 18 (13 – 28) | 1.01 (0.99 - 1.04)<sup>7</sup> | 0.288 |
| **ALP (30-130 IU/L)** | 285 | 78 (63 – 102) | 76 (63 – 99) | 84 (66 – 109) | 1.02 (1.00 - 1.05)<sup>7</sup> | 0.029 |
| **Bilirubin (0-21 µmol/L)** | 296 | 9 (6 – 12) | 9 (7 – 12) | 8 (6 – 14) | 1.02 (0.98 - 1.07) | 0.228 |
| **Albumin (35-50 g/L)** | 298 | 38 (35 – 41) | 39 (36 – 42) | 37 (34 – 40) | 0.90 (0.85 - 0.95) | <0.001 |
| **Heart rate** | 307 | 89 (77 – 105) | 90 (77 – 106) | 90 (79 – 107) | 1.01 (0.95 - 1.07)<sup>7</sup> | 0.745 |
| **Systolic blood pressure** | 307 | 126 (111 – 140) | 125 (111 – 140) | 130 (112 – 140) | 1.03 (0.98 - 1.09)<sup>7</sup> | 0.253 |
| **Diastolic blood pressure** | 307 | 72 (64 – 81) | 74 (66 – 82) | 70 (64 – 78) | 0.94 (0.85 - 1.03)<sup>7</sup> | 0.171 |
| **Respiratory rate** | 306 | 21 (18 – 26) | 20 (18 – 24) | 24 (18 – 30) | 1.09 (1.05 - 1.14) | <0.001 |
| **Oxygen saturations prior to oxygen therapy** | 226 | 95 (91 – 97) | 95 (92 – 97) | 94 (88 – 96) | 0.94 (0.91 - 0.98) | 0.002 |
| **Hypoxia<sup>4</sup>** | 308 | 191/308 (62.0) | 113/207 (54.6) | 60/80 (75.0) | 2.50 (1.42 - 4.52) | 0.002 |
| **Temperature** | 309 | 37.2 (36.6 – 37.9) | 37.2 (36.6 – 37.9) | 37.1 (36.4 – 37.7) | 0.83 (0.62 - 1.09) | 0.182 |
| **Definite COVID-19 on baseline CXR** | 303 | 121/303 (39.9) | 78/198 (39.4) | 31/80 (38.8) | 0.97 (0.57 - 1.65) | 0.921 |
| **CURB65 score<sup>5</sup>** | 299 | 2 (1 – 2) | 1 (0 – 2) | 2 (1 – 3) | 2.31 (1.73 - 3.13) | <0.001 |

Data are median (IQR) for continuous variables, and n (%) (or n/N (%)) for categorical variables. Local laboratory normal ranges for blood tests are shown in parentheses. Multivariate logistic regression was performed in 277 patients with complete data for all variables in the multivariate model. <sup>1</sup>Presence of at least one of respiratory comorbidity, heart failure, diabetes, active cancer or immunosuppression. <sup>2</sup>Defined as at least one of: asthma, COPD, interstitial lung disease, obstructive sleep apnoea, home nebuliser/oxygen/non-invasive pressure support. <sup>3</sup>Defined as at least one of: immunodeficiency syndrome, maintenance steroids (prednisolone ≥ 5mg/day, hydrocortisone ≥ 15mg/day, any dose dexamethasone); conventional synthetic immunosuppressive drugs (excluding hydroxychloroquine and sulfasalazine); biologics; JAK inhibitors; cytotoxic chemotherapy within past 6 months. <sup>4</sup>Defined as oxygen saturations ≤ 94% on room air, or any use of supplemental oxygen. <sup>5</sup>CURB65 with 1 point each for confusion, urea > 7, RR > 30, systolic blood pressure < 90 mmHg or 60 mmHg diastolic, and age ≥ 65. <sup>6</sup>OR stated per 10 unit increase in the independent variable. <sup>7</sup>OR stated per 5 unit increase in the independent variable. <sup>8</sup>OR stated per 0.01 unit increase in arterial blood pH.
ACE: angiotensin converting enzyme, ALP: alkaline phosphatase, ALT alanine aminotransferase, ARB: angiotensin receptor blocker, CRP: C-reactive protein, COPD: chronic obstructive pulmonary disease, CXR: chest x-ray, CRP: C-reactive protein, eGFR: estimated glomerular filtration rate, IQR: interquartile range, N: total number of measurements for each variable for the entire cohort, OR: odds ratio, WBC: white blood cell.

Where baseline oxygen saturations were available, 192/308 (62.3%) patients were hypoxic on admission (≤94% or requiring supplemental oxygen). 174/308 (56.5%) patients presented with COVID-19-associated severe pneumonia according to World Health Organisation criteria (defined as oxygen saturations ≤93% without supplemental oxygen and/or respiratory rate > 30 breaths/min)11.

Oxygen treatment had already been initiated in 83 (26.3%) patients prior to the first recorded set of hospital observations. Where available, the median (IQR) initial oxygen saturation measurement prior to commencement of oxygen therapy was 95% (91 – 97%). Arterial blood gas (ABG) measurements were available prior to oxygen therapy for 97 patients, with a median (IQR) PaO2 of 8.20 (6.50 – 9.40) kPa. A total of 153 patients had ABG on admission (including those receiving supplemental oxygen therapy), revealing a median (IQR) PaO2 of 8.20 (6.40 – 9.70) kPa.

Most patients had elevated acute phase reactants on admission, with median (IQR) [range] C-reactive protein (CRP) of 72 (30 – 131) [<5 – 523] mg/dL and median (IQR) [range] ferritin of 653 (306.5 – 1262.8) [40 – 8685] µg/L, the latter measured in 168 patients. Lymphopaenia (<1 x 109/L) was observed in 186/311 (60.1%) patients, and 246/305 (80.7%) were eosinopaenic (<0.04 x 109/L). In those who underwent testing, elevations were noted in lactate dehydrogenase (94/116, 81.0%), D-dimer (19/33, 57.6%), troponin-I (28/52, 53.8%) and creatine kinase (48/141, 34.0%) (Table 1).

Chest X ray (CXR) was performed on admission for 303 (95.9%) patients. Of the 13 patients without a CXR, 12 had mild disease with no significant lower respiratory tract symptoms and one had severe disease in the context of active malignancy and received palliative care from admission. 196/303 (64.7%) CXRs were abnormal at baseline with 121/303 (39.9%) reported as ‘classic/probable’ COVID-19 according to the British Society of Thoracic Imaging reporting template.9 Of those with normal initial CXR, 31/107 (29.0%) patients had abnormal radiology on repeat imaging. Computed tomography (CT) of the chest, though not part of routine COVID-19 investigations in our Trust, was performed on clinical discretion in 40 patients, of which 27 were diagnostic of COVID-19. Overall, 227/303 (74.9%) of SARS-CoV-2 PCR positive patients had abnormal thoracic imaging.

### Patient outcomes and management decisions

Daily clinical status (including survival post-discharge) was recorded for a median (IQR) follow-up duration of 28 (20 – 28) days after admission. Re-admission to hospital occurred in 27/316 (8.5%) patients, of whom 7 died. In total, 81 (25.6%) patients died, 210 (66.5%) were discharged and remained alive at the point of analysis and 25 (7.9%) required ongoing inpatient care. Several nonrespiratory complications of COVID-19 were noted. The most common associated diseases were cardiac dysrhythmias (22 [7%]), heart failure (11 [3.5%]), and enterocolitis (10 [3.2%]). 18/316 (5.7%) patients developed vascular complications including stroke (9 [2.9%]), pulmonary embolus (6 [1.9%]), and limb ischaemia (3 [1%]). Antibiotics were prescribed for 255 (80.7%) patients. 60/316 (19.0%) patients were admitted to critical care with a median (IQR) [range] duration of intensive care unit (ICU) admission of 8.5 (3 – 15) [1–34] days. Within ICU, vasopressors and renal replacement therapy were required for 28/60 (46.7%) and 9/60 (15.0%) patients respectively. An epidemic curve showing the daily incidence of admissions, ventilatory support, and mortality for the entire cohort is shown in Figure 1.

**Figure 1.**
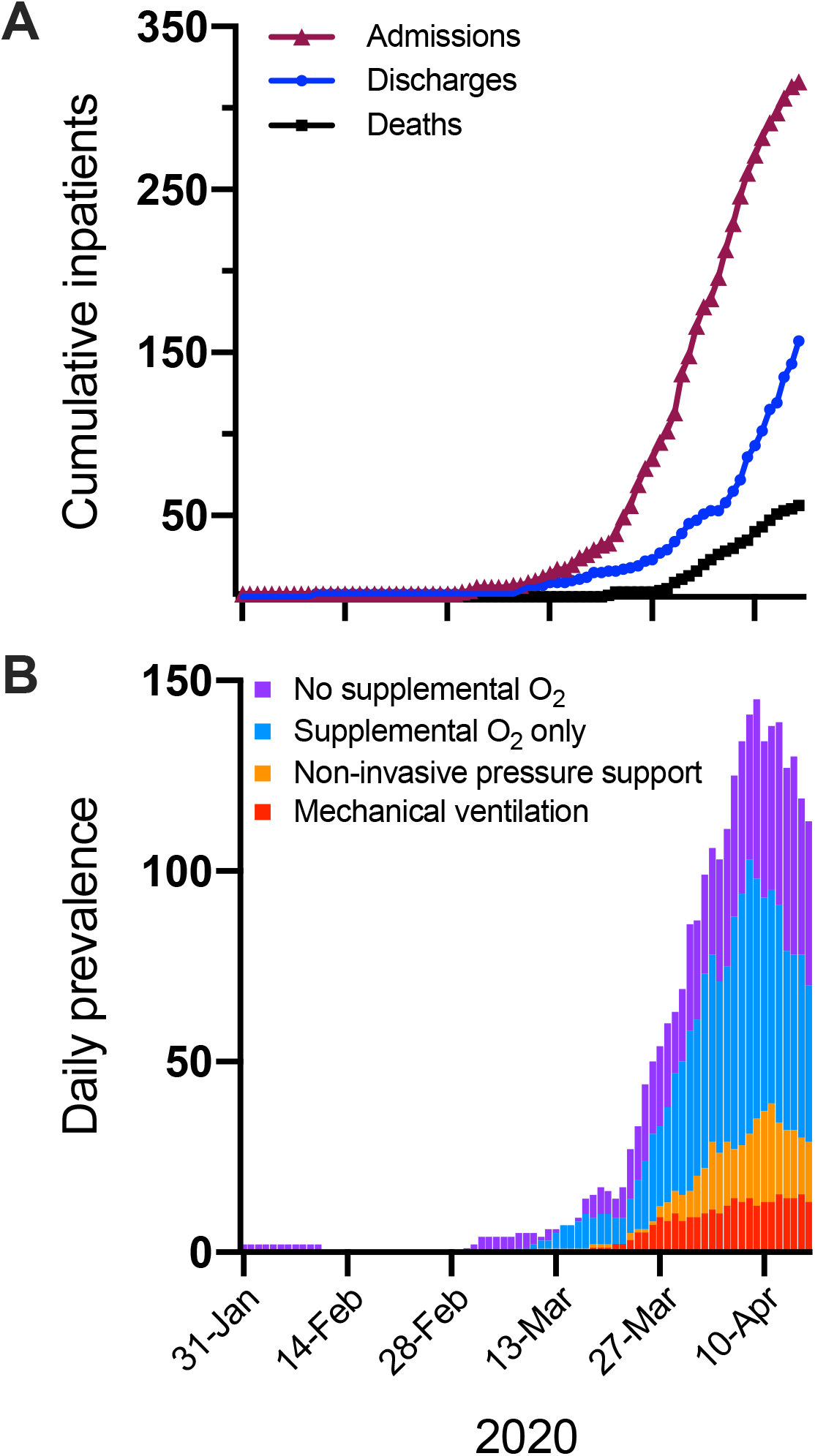
A: Cumulative daily incidence of admissions, discharges and deaths up to censor point of16th April 2020. B: Daily prevalence of inpatients with COVID-19 by oxygen and ventilation requirements up to censor point of 16th April 2020.

We ascertained 28-day outcome data (death, or discharged and remains alive) for 291/316 (92.1%) patients (the other 25 remained inpatients at the time of analysis). The median (IQR) duration of hospital admission was 8 (4 – 12) days and 81/291 (27.8%) patients died. 223 patients did not receive ventilation, of which 171/223 (76.7%) were alive beyond discharge (Figure 2). Three patients died in the community after discharge, two of whom were discharged with end of life care plans, while one was reviewed by their general practitioner and received end of life care in the community.

**Figure 2.**
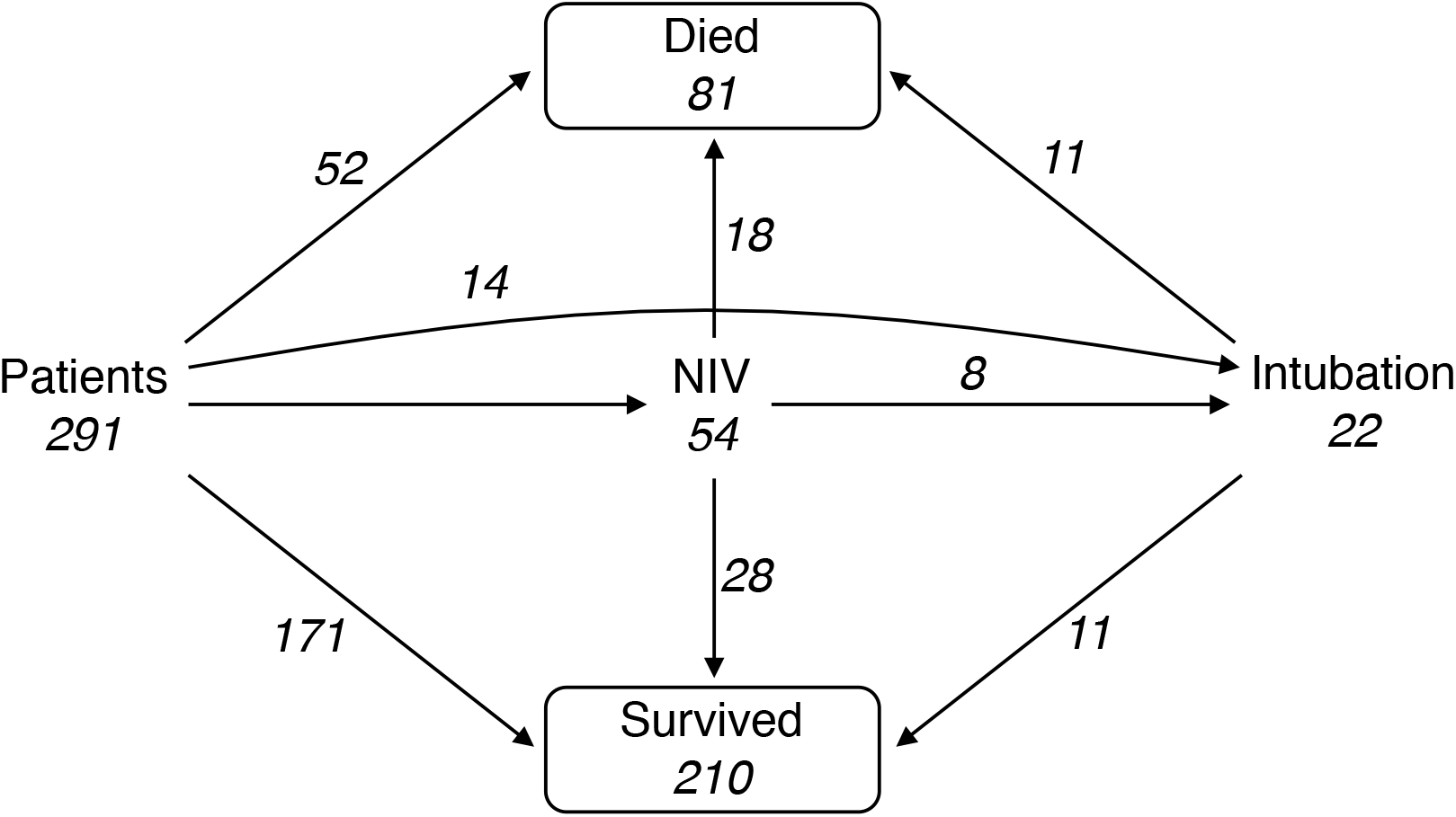
Schematic summarising interventions and outcomes for 291 inpatients with a confirmedclinical outcome. NIPS: non-invasive pressure support.

Of the 52 patients who died in hospital without receiving ventilatory support, all deaths were expected and occurred after an advanced decision regarding ceiling of treatment was reached with the patient (where possible) and/or their relatives. Patients who died without ventilatory support were generally frail with a median (IQR) Clinical Frailty Scale score of 7 (6 – 7), and compared to the rest of the cohort were significantly older (median [IQR] age: 85 [79-91] vs. 71 [56-80], p < 0.001) and more likely to live in a nursing/residential home (33/52 [63.5%] vs. 21/239 [8.8%], p < 0.001). Of these patients, 44/49 (89.9%) had anticipatory palliative care medications prescribed and 43/49 (87.8%) were reviewed by a member of the hospital specialist palliative care team before death. Close relatives could visit prior to death with appropriate personal protective equipment in line with our end of life infection control policy.

54 patients received non-invasive pressure support (NIPS) (Figure 2), of which 51 received continuous positive airway pressure and three received bilevel positive airway pressure. Of these, 29 patients received NIPS as their ceiling of treatment, 17 had NIPS without a need for mechanical ventilation, and eight were subsequently intubated and mechanically ventilated for progressive respiratory failure. An additional 14 patients received immediate mechanical ventilation without prior NIPS. Half of patients who received either NIPS (28/54) or mechanical ventilation (11/22) survived beyond discharge. Two thirds (18/29 [62.1%]) patients who received NIPS as their ceiling of treatment died. By contrast, in those receiving NIPS as part of a plan to escalate to intubation where necessary, two thirds (17/25 [65.3%]) survived.

46/291 (15.8%) patients were admitted to ICU, of which 13/46 (28.3%) died. Of these, 8/46 required only supplemental oxygen (all survived), 16 (34.8%) were escalated to NIPS only (mortality 2/16 [12.5%]), and 22 (47.8%) were intubated and mechanically ventilated (mortality 11/22 [50%]).

### Association of baseline factors with mortality

We explored the association between baseline factors measured at the point of admission with mortality by univariate logistic regression (Table 1). Increasing age was strongly associated with mortality (Figure 3), with a median age of 82 years in those who died versus 68 years in those who survived (OR 1.09 per year [1.06 – 1.12], p < 0.001). A shorter symptom duration was also associated with increased mortality (median 3 vs. 6 days, OR 0.92 [0.86 – 0.97], p = 0.004), as was pre-existing heart failure (OR 2.67 [1.36 – 5.19], p = 0.004), hypertension (OR 1.95 [1.16 – 3.3], p=0.011) and dementia (OR 3.50 [1.87 – 6.58], p < 0.001). In keeping with the main presenting feature of COVID-19, hypoxia on admission (OR 2.50 [1.42 – 4.52], p = 0.002) and raised respiratory rate (1.09 [1.05 – 1.14] per breath/min increase, p<0.001) were associated with death. Reduced renal function was also associated with increased mortality (estimated glomerular filtration rate (eGFR): OR 0.97 [0.96 – 0.98] per 5 mL/min.1.73m2 increase, p<0.001; urea: OR 1.11 [1.06 – 1.16] per mmol/L increase, p<0.001), as was elevated CRP (OR 1.04 (1.01 – 1.07) per 10mg/L increase, p = 0.005). The presence of severe COVID-19 pneumonia,11 and higher CURB65 score,12 were both associated with death.

**Figure 3.**
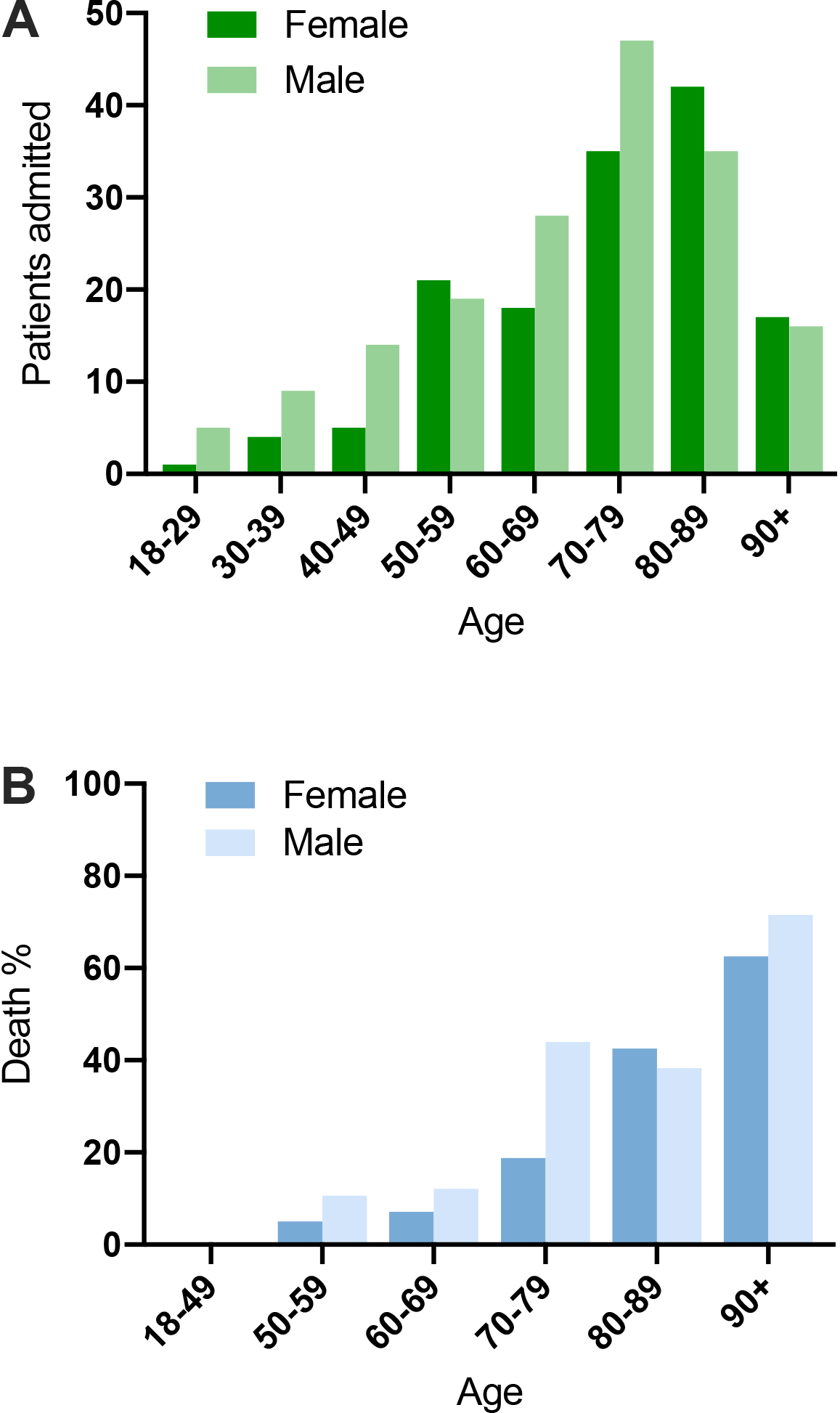
A: Admissions by age and sex of cohort of 316 patients. B: Percentage of deaths among 291patients with a confirmed clinical outcome, stratified by age and sex.

Based on univariate analyses, we did not identify any significant effect of pre-admission use of angiotensin-converting enzyme (ACE) inhibitor or angiotensin receptor blocker (ARB) use with death (OR 0.78 [0.41 – 1.43], p = 0.43). Furthermore, there was no significant association between mortality and the presence of diabetes, immunosuppression, or any respiratory comorbidity, though this may reflect the smaller sample size. Although lymphopaenia was frequently observed, the absolute lymphocyte count was not significantly associated with mortality (OR 0.99 [0.87 – 1.07] per 1 x 109/L increase, p = 0.85).

We selected variables to include as prospective candidates in a multiple logistic regression satisfying all of the following criteria: unambiguous to obtain from retrospective data, demonstrating a clinically meaningful difference, unlikely (based on clinical knowledge) to be a proxy measure for another variable in the dataset, and with sufficient available baseline data. Based on these criteria, we selected seven key variables: age, male sex, WHO severity classification for severe pneumonia,^11^ CRP, eGFR, heart failure, and hypertension. We performed multiple logistic regression analyses for patients with complete data for all variables (n = 274) exploring the sensitivity of the solutions to the inclusion or exclusion of other candidates. The only variables withstanding robustness testing were age (OR 1.09 [1.06 – 1.12] per year increase, p < 0.001) and WHO severity at presentation (OR 2.54 [1.38 – 4.67], p = 0.002). In addition, there was some evidence of an interaction between these two variables (p = 0.022): mortality was higher in older patients regardless of severity; death also occurred in younger patients, but only in those with high severity at presentation.

## Discussion

Outcomes in our cohort broadly reflect national UK experience. Crude mortality was 25.6%, compared to 33% in the ISARIC cohort.^6^ After restricting analysis only to those with a definite outcome at the point of analysis (minimising bias towards reduced deaths), our overall mortality rate was 27.8%, compared to similar numbers in US (21%)^5^ and China (28%)^3^ datasets and 40% in ISARIC.^6^ There are many possible factors that might influence differences in crude mortality rates between and within countries, including admission policy, demographics, disease severity in those admitted, testing criteria and inpatient management. The ISARIC data reveal an older population of hospitalised patients in the UK compared to other countries, with a greater burden of comorbidity and imply the possibility of advanced decision making by clinicians; however, granular detail was lacking.

Our data shed light on the clinical decisions occurring both in general wards and in critical care settings. Whilst admission to critical care in our cohort (18.9%) was comparable to national (17%)^6^–and international (7.5-26%)^3–5^ experience, there was also clear evidence of advanced decision making for individual patients. In all patients who died without receiving ventilation, a ceiling of treatment plan was discussed in advance with the patient and/or their family and documented in the medical record. These patients were more frail, had a higher number of comorbidities, and were more likely to reside in a residential or nursing home, implying that their premorbid risk of death was high. Similarly, a third of patients who died received NIPS for single organ (respiratory) failure where there was an advance decision not to escalate to mechanical ventilation in the event of a failure to respond. Thus death was anticipated in most patients dying in hospital, and palliative care teams provided specialist input into patient management. Integral to our patient-centred approach was to implement a policy of permitting a single visit from relatives, using personal protective equipment, to patients at the end of life.

Death occurred in 11 patients out of 22 receiving mechanical ventilation (50%), matching the UK experience from ISARIC (53%).6 These outcomes compare favourably with early international reports of extremely high rates of death (97%) in ventilated patients.3 Rates of mechanical ventilation were 497/6628 (7.5%) in the ISARIC cohort and 22/316 (6.9%) in our cohort, which appears lower than in the US (20%)5 and China (17%).^3^ A possible interpretation is that mechanical ventilation has been applied more selectively in the UK to a hospitalised population with high rates of frailty and comorbidity – an approach that is consistent with national guidance for critical care management of patients with COVID-19.13 In our cohort, NIPS was widely used, either as the ceiling of treatment in patients with respiratory failure and for whom escalation to mechanical ventilation was not considered appropriate (Burns et al., submitted), or as a bridge to mechanical ventilation in critical care and on medical wards. The possibility that NIPS is of benefit in management of COVID-19, and the optimal patient group(s) and timing of initiation is being addressed in clinical trials.

Baseline characteristics of our cohort reflect the broader UK experience. Our cohort had a median age of 75 and included very few patients under 40 (6%). Most patients had one or more comorbidities and approximately one fifth of our cohort lived in a care home. Presenting symptoms were also consistent with national and international cohorts. Nevertheless, there were also differences. Nosocomial infections made up a low proportion of total cases (21/362 [5.8%]) and relatively few patients were healthcare workers (27/316 [8.5%]) or people of Black, Asian and minority ethnic (BAME) background (7.3%). Eight patients were admitted when management of COVID-19 occurred exclusively in HCID units in order to prevent community spread; these patients were younger, with mild disease, and all survived.

Unlike other countries but consistent with UK practice, CT was infrequently used to diagnose COVID-19 in our setting, whereas CXR was widely used. Our data show that the admission CXR was abnormal in around two thirds (64.7%) of patients with COVID-19. This is in line with data from a Hong Kong study of COVID-19 patients where 69% of 255 baseline chest radiographs were abnormal.14 To our knowledge there are no published data on the distribution of COVID-19 features on admission CXRs according to the British Society of Thoracic Imaging reporting template: in our cohort, 39.9% of CXRs were reported as showing ‘classic/probable’ COVID-19.9

Multiple logistic regression of baseline clinical factors that were associated with death highlighted age and disease severity (a composite of oxygen saturations and respiratory rate) as statistically significant factors: of these, age had the more significant association. This is a consistent finding in COVID-19 across the world,^3,15,16^ confirming that regional demographic variations are likely to have a major impact on mortality. Strikingly, we observed no deaths in patients under 55, although overall numbers in this group were relatively small (50/291 [17.2%]). We also examined the influence of ethnicity, finding no evidence of an association. Since there are fewer people from BAME backgrounds in the North East of England compared to other regions, nothing can be inferred from these findings. We were unable to analyse the influence of obesity, which was identified as a factor in the ISARIC cohort,^6^due to missing data on height and weight in this acutely unwell cohort (we considered reports of obesity in clinical notes to be unreliable). Interestingly, laboratory values, including CRP or lymphocyte count, did not predict outcome, although our data were underpowered to detect differences in these factors.

This study has several limitations. Data were retrospectively collected at a single NHS Trust, and may therefore not reflect COVID-19 transmission patterns in other parts of the UK nor necessarily reflect inpatient management across the wider NHS. Whilst our cohort size is similar to published analyses,^3,17^ the number of patients is relatively low. In addition, the modelling was based on a subset of patients for which adequate data was available and excluded those with nosocomial infection. Strengths of this analysis are the extended length of follow up, which is longer than most published cohorts, the large proportion of cases with definite clinical endpoints, and robust clinical informatics mechanisms to capture deaths in the community occurring after discharge.

This report is the first to provide a detailed description of the inpatient management of COVID-19 at the individual patient level, complementing and enriching existing literature. These results will be broadly informative to clinicians, policy makers and healthcare providers.

## Contribution statement

KFB and CJAD conceived the study. KFB, EH and CJAD designed the data collection plan. NUTH COVID Control and Clinical Groups, LP-C, AW, BAIP, YT, DAP, CG, MLS, EH, CJAD provided clinical care and/or developed patient pathways. KFB, ATH, ISvdL, SAT, RC, GM, AL, AB, AE, SA, DB, OM, NA and JF collected data. KFB, ATH, ISvdL, CL, DL, EH, and CJAD analysed and interpreted data. KFB, CL, and DL performed statistical modelling. KFB, ATH, ISvdL, and CJAD drafted the manuscript. All authors reviewed the draft version of the manuscript and approved the final version for publication. The corresponding author had full access to all data and takes responsibility for the decision to submit for publication.

## Data Availability

Clinical data protected by Caldicott regulations

## Declaration of interests

The authors declare no competing interests.

## Acknowledgements

CJAD is funded by the Wellcome Trust (211153/Z/18/Z). KFB is funded by a National Institute for Health Research (NIHR) Clinical Lectureship (CL-2017-01-004). BAIP is funded by the Wellcome Trust (109975/Z/15/Z, 203105/Z/16/Z). It is impossible to individually acknowledge all of those involved, however we express our deep gratitude to all colleagues within the Newcastle upon Tyne NHS Foundation Trust, including but not exclusively medical, nursing, allied health professional, laboratory, support and non-clinical staff, for their enormous collective effort in delivering the patient care described herein. The views expressed are those of the authors and not necessarily those of the NHS, the NIHR or the Department of Health and Social Care. The funders had no role in study design, data collection, or decision to publish.

## Notes

### Competing Interest Statement

The authors have declared no competing interest.

### Funding Statement

No funding was required

